# Serum IgA and IgM levels in hemochromatosis probands with *HFE* p.C282Y homozygosity

**DOI:** 10.1101/2025.06.23.25330000

**Authors:** James C. Barton, J. Clayborn Barton, Luigi F. Bertoli, Ronald T. Acton

## Abstract

**Background:** Serum IgA and IgM levels in cohorts of adults with hemochromatosis are not reported.

**Methods:** We compiled serum IgA and IgM levels at diagnosis of hemochromatosis in probands with *HFE* p.C282Y (rs1800562) homozygosity, investigated associations of IgA and IgM with clinical and other laboratory characteristics, and compared mean IgA and IgM of probands with combined/weighted means of published adult European cohorts not selected for hemochromatosis.

**Results:** There were 73 probands (36 men, 37 women; mean age 51 ± 13 y). Fifty probands (68.5%) had human leukocyte antigen (HLA)-A*03. Mean IgA ± standard deviation [95% confidence interval] was 2.11 ± 1.06 g/L [1.87, 2.35]. Mean IgM was 1.11 ± 0.75 g/L [0.94, 1.28]. IgM was inversely associated with age (Pearson’s r_73_ = –0.2733; p = 0.019). A multiple regression on IgA revealed no significant association with other characteristics. A regression on IgM revealed one positive association (daily alcohol intake; p = 0.036) and one negative association (age; p = 0.016). Mean IgA of male and female probands and corresponding mean IgA of Europeans in two cohorts (918 men, 458 women) did not differ significantly. Mean IgM of probands was lower than the mean IgM of Europeans in four cohorts (men 1.03 ± 0.84 g/L vs. 1.35 ± 0.55 g/L (n = 1084)), respectively (p <0.001); women 1.18 ± 0.67 g/L vs. 1.57 ± 0.68 g/L (n = 622), respectively (p <0.001)).

**Conclusions:** Serum IgM levels of hemochromatosis probands with *HFE* p.C282Y homozygosity are positively associated with daily alcohol intake and inversely associated with age. Mean IgM levels of male and female probands are lower than those of European men and women not selected for hemochromatosis.

## Introduction

Hemochromatosis in persons of western European descent is associated with homozygosity for *HFE* p.C282Y (rs1800562), a common missense allele of the homeostatic iron regulator (chromosome 6p22.2) in linkage disequilibrium with human leukocyte antigen (HLA)-A*03 [1,2]. HFE, a non-classical class I major histocompatibility complex protein, is an upstream regulator of hepcidin and thus of iron homeostasis [3]. The estimated prevalence of p.C282Y homozygotes in newborns in Ireland is 1 in 100 [4], in persons of European descent in the United Kingdom is 1 in 156 [5], and in non-Hispanic white adults in North America is 1 in 227 [6].

Laboratory phenotypes of many adults at diagnosis of *HFE* p.C282Y homozygosity include elevated transferrin saturation (TS) and serum ferritin (SF) [7]. Adults with p.C282Y homozygosity have increased risks of developing iron overload that may contribute to the development of arthropathy, diabetes mellitus, cirrhosis, hypogonadotropic hypogonadism, or cardiomyopathy [7]. Severe iron overload occurs predominantly in men [7,8]. Non-*HFE* heritable and environmental variables modify iron loading in adults with p.C282Y homozygosity [2,7,9–11].

*HFE* p.C282Y homozygotes in Denmark, including those with normal TS and SF, had greater risks of any infection, sepsis, and death from infections than persons with other *HFE* genotypes [12]. Studies of persons not selected for hemochromatosis diagnoses or *HFE* genotypes suggest that lower levels of immunoglobulin A (IgA) [13] and immunoglobulin M (IgM) [14] contribute to the occurrence or severity of infections, including sepsis, although we found no reports of serum IgA and IgM levels in cohorts of adults with p.C282Y homozygosity.

The aims of this retrospective study were: 1) to compile serum IgA and IgM levels at diagnosis in 73 referred hemochromatosis probands with *HFE* p.C282Y homozygosity; 2) to determine associations between serum IgA and IgM levels and clinical and other laboratory characteristics at diagnosis of hemochromatosis; and 3) to compare mean serum IgA and IgM levels of this cohort with combined/weighted mean serum IgA and IgM levels from published cohorts of European adults not selected for hemochromatosis diagnoses or *HFE* genotypes.

## Methods

### Ethics statement

This retrospective work was performed according to the principles of the Declaration of Helsinki [15]. The performance of this study was approved by the Western Institutional Review Board, Inc. (submission 2539985-44189619). Western Institutional Review Board, Inc. waived the need for obtaining informed consent from participants in this study under the United States Department of Health and Human Services, Office for Human Research Participants, regulation 45 CFR 46.101(b)(4). Informed consent was not required and thus was not obtained because this study involved retrospective chart reviews and analyses of observations recorded in routine medical care.

Data analyzed in this study were not anonymized before the investigators accessed them because data were compiled from proband charts in an Alabama tertiary hematology center wherein JaCB and LFB diagnosed and treated all probands, consistent with Western Institutional Review Board, Inc. approval of this study. JaCB, JClB, and LFB had access to information that could identify individual probands during and after data collection. Data were compiled and analyzed during the interval 30 December 2018 - 3 June 2020. All data in this report are displayed in a manner that maintains proband anonymity in both the present results and the corresponding dataset [16].

### Definition of *HFE*-related hemochromatosis

We defined *HFE*-related hemochromatosis as p.C282Y homozygosity, the current diagnostic criterion promulgated by an expert group of the BIOIRON Society [17]. Because it is widely recognized that the penetrance of high-iron phenotypes in p.C282Y homozygotes is low [6,17], we included subjects with p.C282Y homozygosity regardless of their iron phenotypes.

### Subjects included

We retrospectively compiled data on all consecutive patients aged ≥18 y referred to an Alabama tertiary hematology center during the study interval 1 January 2007 - 30 October 2018 for evaluation and management of hemochromatosis who met the following criteria: a) had *HFE* p.C282Y homozygosity, b) had no known non-hemochromatosis iron disorder, c) underwent measurements of serum IgA and IgM at diagnosis, d) achieved iron depletion with therapeutic phlebotomy, as appropriate, and e) were the first in their respective families to be diagnosed to have hemochromatosis (probands). Alabama hemochromatosis probands with p.C282Y homozygosity previously reported 82.5% British Isles ancestry [18].

We evaluated the medical records of 169 referred hemochromatosis probands with *HFE* p.C282Y homozygosity and excluded 12 probands (three with viral hepatitis B or C, three with polyclonal gammopathy, two with monoclonal gammopathy, and four without sufficient data). Of the remaining 157 probands, we included the 73 probands (46.5%) in the present study for whom IgA and IgM data were available and who met other criteria for inclusion.

Medical histories were taken from probands and the records of referring physicians. All probands underwent medication review, physical examination, laboratory testing, imaging procedures, and evaluation of these iron overload-related conditions (hemochromatosis hand arthropathy, diabetes, hypogonadotropic hypogonadism, cardiomyopathy, and cirrhosis) at diagnosis, as appropriate, and as described in detail elsewhere [19,20]. We defined obesity as BMI ≥30 kg/m^2^ [21]. We classified proband reports of alcohol intake as a dichotomous variable: 1) daily alcohol intake or 2) infrequent or no alcohol intake. We reviewed medical charts of probands who had either subnormal serum IgA or subnormal serum IgM and compiled their reports of frequent, severe, or unusual infections.

### Subjects excluded

We excluded subjects with the following characteristics: hyperferritinemia, hemochromatosis, or *HFE* p.C282Y homozygosity diagnosed as a consequence of family or population screening; diagnosis of a primary or secondary hematologic disorder; volunteer donation of more than two units of whole blood in the year before hemochromatosis diagnosis; bariatric operation [22]; viral hepatitis B or C; liver transplant; diagnosis of malignancy; anti-cancer therapy; non-iron-related chronic inflammatory condition; self-reported pregnancy; monoclonal or polyclonal gammopathy; previous diagnosis of primary antibody deficiency; or IgA nephropathy.

We excluded probands treated with the following drugs that have been reported to decrease IgA levels: agents used to treat inflammatory bowel disease [23]; anti-seizure agents [24–26]; captopril [27]; cyclosporine [28]; d-penicillamine [29,30]; fenclofenac [31]; gold compounds [32]; hydroxychloroquine [33]; methotrexate [34]; oral corticosteroids [35]; rituximab [36]; and sulfasalazine [37]. We also excluded probands treated with the following drugs that have been reported to decrease IgM levels: agents used to treat inflammatory bowel disease [23]; anti-seizure agents [24–26]; diclofenac and fenclofenac [38]; d-penicillamine [30]; levamisole [39]; methotrexate [34]; natalizumab [40]; and rituximab [36].

### Laboratory

Blood specimens were collected in the mornings without regard to fasting. Complete blood counts were measured using an automated hematology analyzer (Cell-Dyn® Model 610, Model 1700, Model 1800, or Emerald (Abbott Laboratories, Chicago, IL, USA)) within one hour after specimen collection. The reference range for absolute lymphocyte count was the same for each analyzer (0.6-4.1 x 10^6^/L). TS and SF were measured using standard clinical laboratory methods (Laboratory Corporation of America, Burlington, NC, USA). We defined these TS and SF levels to be elevated: TS >50% (men) and TS >45% (women); and SF >300 µg/L (men) and SF >200 µg/L (women) [6,41].

Serum IgA and IgM were measured using rate nephelometry (Laboratory Corporation of America, Burlington, NC, USA). The following reference limits were based on 1996 consensus guidelines [42]: IgA 0.91-4.14 g/L (91-414 mg/dL) and IgM 0.40-2.30 g/L (40-230 mg/dL). We defined elevated IgA as levels >4.14 g/L and subnormal IgA as levels <0.91 g/L. We defined elevated IgM as levels >2.30 g/L and subnormal IgM as levels <0.40 g/L. We did not measure IgA subclasses or serum kappa or lambda light chains.

*HFE* genotyping was performed as previously described [19]. We determined HLA-A types as previously described [19,43] and defined HLA-A*03 as the marker for the hemochromatosis ancestral haplotype [19,44,45]. Positivity for HLA-A*03 was defined as either homozygosity or heterozygosity.

### Phlebotomy units removed to achieve iron depletion

Iron depletion therapy, defined as the periodic removal of blood to eliminate storage iron, was performed in probands with elevated SF levels as described elsewhere [46]. We defined 450-500 mL of blood removed at a single session as one phlebotomy unit. Iron depletion therapy was complete when SF was less than 20 μg/L [46]. We defined the number of phlebotomy units removed to achieve iron depletion in probands without elevated SF as zero.

### Serum IgA and IgM in published European cohorts

We performed computerized and manual searches to identify a convenience sample of published reports of serum IgA and IgM measured in cohorts of more than 50 healthy, “control,” or general population European adults aged ≥18 y not selected for hemochromatosis diagnoses or *HFE* genotypes. We selected reports that described laboratory methodology for measurement of serum IgA and IgM and displayed Ig cohort data as means, standard deviations (SD), and 95% confidence intervals (CI). We converted IgA and IgM measures published as IU/L to g/L according to the method of Rowe et al. [47]. We excluded reports in which serum IgA and IgM measurements were expressed only as medians.

### Statistics

The dataset for analysis is displayed in a Figshare file [16]. There were observations on 73 probands (36 men, 37 women). Kolmogorov-Smirnov testing demonstrated that age, BMI, IgA, and IgM data did not differ significantly from those that are normally distributed. We displayed these data as means ± 1 SD and compared them using the Student’s t test for unpaired data (two-tailed). TS, SF, and phlebotomy unit data differed significantly from those that are normally distributed. We displayed these data as medians (ranges) and compared them using the Mann-Whitney U test (two-tailed). Categorical variables were compared using Fisher’s exact test (two-tailed).

Frequency distributions of IgA and IgM, displayed as smoothed curves, depict percentages of 73 probands as functions of ten subgroups of the corresponding Ig levels. Error bars represent 95% CI with continuity corrections for the proportions of probands in each subgroup.

We evaluated these independent variables for suitability in multiple regressions on serum IgA and IgM: age, sex, daily alcohol intake (dichotomous), hemochromatosis arthropathy, diabetes, cirrhosis, BMI, TS, SF, absolute lymphocyte count, IgA, IgM, and positivity for HLA-A*03. A preliminary regression model on IgA revealed that standardized beta coefficients (beta) were low and values of p were high for all independent variables except daily alcohol intake, hemochromatosis arthropathy, and TS, and thus we excluded these insignificant variables from the final IgA regression model. A preliminary regression model on IgM revealed that standardized beta coefficients (beta) were low and values of p were high for all independent variables except age and SF, and thus we excluded these insignificant variables from final IgM regression model.

We used a Cochrane formula [48] to combine the Ig-specific n, mean, and SD from multiple published cohorts into descriptors of a single group. We used a two-sample t test and n, mean, and SD from each group to compare Alabama proband data with those of Europeans not selected for hemochromatosis diagnoses or *HFE* genotypes (S1 Tables).

We used Excel^®^ 2000 (Microsoft Corp., Redmond, WA, USA) and GraphPad Prism 8^®^ (2018; GraphPad Software, San Diego, CA, USA). We defined p <0.05 to be significant.

## Results

### Characteristics of probands

There were 36 men (49.3%) and 37 women (50.7%) (Table 1). The mean age of 73 probands was 51 ± 13 y (range 22–80). The prevalence of reports of daily alcohol intake, BMI, TS, SF, and phlebotomy units to achieve iron depletion were greater in men than in women (Table 1). TS was elevated in 33 of 36 men (91.7%) and 33 of 37 women (89.2%) (p = ∼1.000). SF was elevated in 31 men (86.1%) and 30 women (81.1%) (p = 0.757).

**Table 1.** Referred hemochromatosis probands with *HFE* p.C282Y homozygosity.

| Characteristic | Men (n = 36) | Women (n = 37) | Value of p |
| --- | --- | --- | --- |
| Mean age at diagnosis, y ± SD | 50 ± 13 | 52 ± 13 | 0.619 |
| Reports of daily alcohol intake, % (n) | 38.9 (14) | 16.2 (6) | 0.038 |
| Mean body mass index, kg/m<sup>2</sup> ± SD | 30.0 ± 5.2 | 25.9 ± 6.4 | 0.004 |
| Hemochromatosis arthropathy, % (n) | 16.7 (6) | 21.6 (8) | 0.768 |
| Diabetes, % (n) | 19.4 (7) | 5.4 (2) | 0.085 |
| Cirrhosis, % (n) | 16.7 (6) | 2.7 (1) | 0.056 |
| Median transferrin saturation, % (range)<sup>a</sup> | 84 (41, 100) | 73 (21, 100) | 0.020 |
| Median serum ferritin, μg/L (range) | 941 (71, 5630) | 406 (32, 5000) | <0.001 |
| Median absolute lymphocyte count $\times 10^6$/L (range) | 2.0 (1.1, 4.8) | 2.1 (1.0, 3.6) | 0.889 |
| HLA-A*03 positivity, % (n)<sup>b</sup> | 77.8 (28) | 59.5 (22) | 0.131 |
| Mean serum IgA, g/L ± SD | 2.19 ± 0.91 | 2.04 ± 1.20 | 0.538 |
| Mean serum IgM, g/L ± SD | 1.03 ± 0.84 | 1.18 ± 0.67 | 0.402 |
| Median phlebotomy units to achieve iron depletion (range)<sup>c</sup> | 16 (0, 100) | 8 (0, 75) | 0.016 |

*HFE*, homeostatic iron regulator; HLA, human leukocyte antigen; IgA, immunoglobulin A; IgM, immunoglobulin M; SD, standard deviation. No proband had hypogonadotropic hypogonadism or cardiomyopathy attributed to iron overload. All characteristics were determined at diagnosis except phlebotomy units to achieve iron depletion.
<sup>a</sup> Observations in 35 men and 37 women.
<sup>b</sup> Heterozygosity or homozygosity.
<sup>c</sup> Phlebotomy units data were available for 31 men and 30 women.

Twelve probands (5 men, 7 women) did not have elevated SF levels at diagnosis. These probands were diagnosed to have *HFE* p.C282Y homozygosity as a consequence of evaluations for: elevated serum levels of hepatic transaminases (n = 3); macrocytosis (n = 3); menorrhagia (n = 2); blood loss due to colonic polyps or carcinoma (n = 2), respectively; multiple autoimmune disorders (n = 1); and lethargy (n = 1).

HLA-A*03 positivity was detected in 50 probands (68.5%). HLA typing as part of paternity testing of 1,318 apparently normal, unrelated white adults from Alabama revealed that 361 had A*03 positivity (27.4%) [43]. The odds ratio for A*03 in the present probands vs. apparently normal, unrelated white adults from Alabama was 5.8 (p <0.001).

The mean serum IgA at diagnosis of 73 probands was 2.11 ± 1.06 g/L. One man and five women had subnormal IgA (2.8% vs. 13.5%, respectively; p = 0.199). A woman in her 50s had frequent sinusitis, bronchitis, and pneumonia and subnormal total IgG/IgG1/IgG2/IgG4/IgA. Five other probands with subnormal IgA did not report frequent, severe, or unusual infections. The lowest IgA was 0.14 g/L. None of the present probands had severe IgA deficiency, as defined by the World Health Organization [49]. One man and two women had elevated IgA (2.8% vs. 5.4%, respectively; p = ∼1.000). The mean IgA in probands with and without HLA-A*03 positivity did not differ significantly (2.15 ± 1.04 g/L vs. 2.03 ± 1.14 g/L, respectively; p = 0.666).

The mean serum IgM at diagnosis of 73 probands was 1.11 ± 0.75 g/L. Four men and two women had subnormal IgM (11.1% vs. 5.4%, respectively; p = 0.430). A woman in her 50s had frequent sinusitis, bronchitis, and pneumonia and subnormal total IgG/IgG1/IgG3/IgG4/IgM. Five other probands with subnormal IgM did not report frequent, severe, or unusual infections. The lowest IgM was 0.11 g/L. None of the present probands had severe IgM deficiency, defined as serum levels less than 10% of the mean IgM [50] of the present cohort. One man and four women had elevated IgM (2.8% vs. 10.5%, respectively; p = 0.358). The mean IgM in probands with and without HLA-A*03 positivity did not differ significantly (1.15 ± 0.84 g/L vs. 1.03 ± 0.52 g/L, respectively; p = 0.540).

### Frequency distribution of groups of serum IgA levels

The frequency distribution of groups of IgA levels was skewed to the right (Fig 1). The range 1.29-2.76 g/L included 67.1% of levels. The range of IgA levels was 0.14-6.05 g/L.

**Figure 1.**
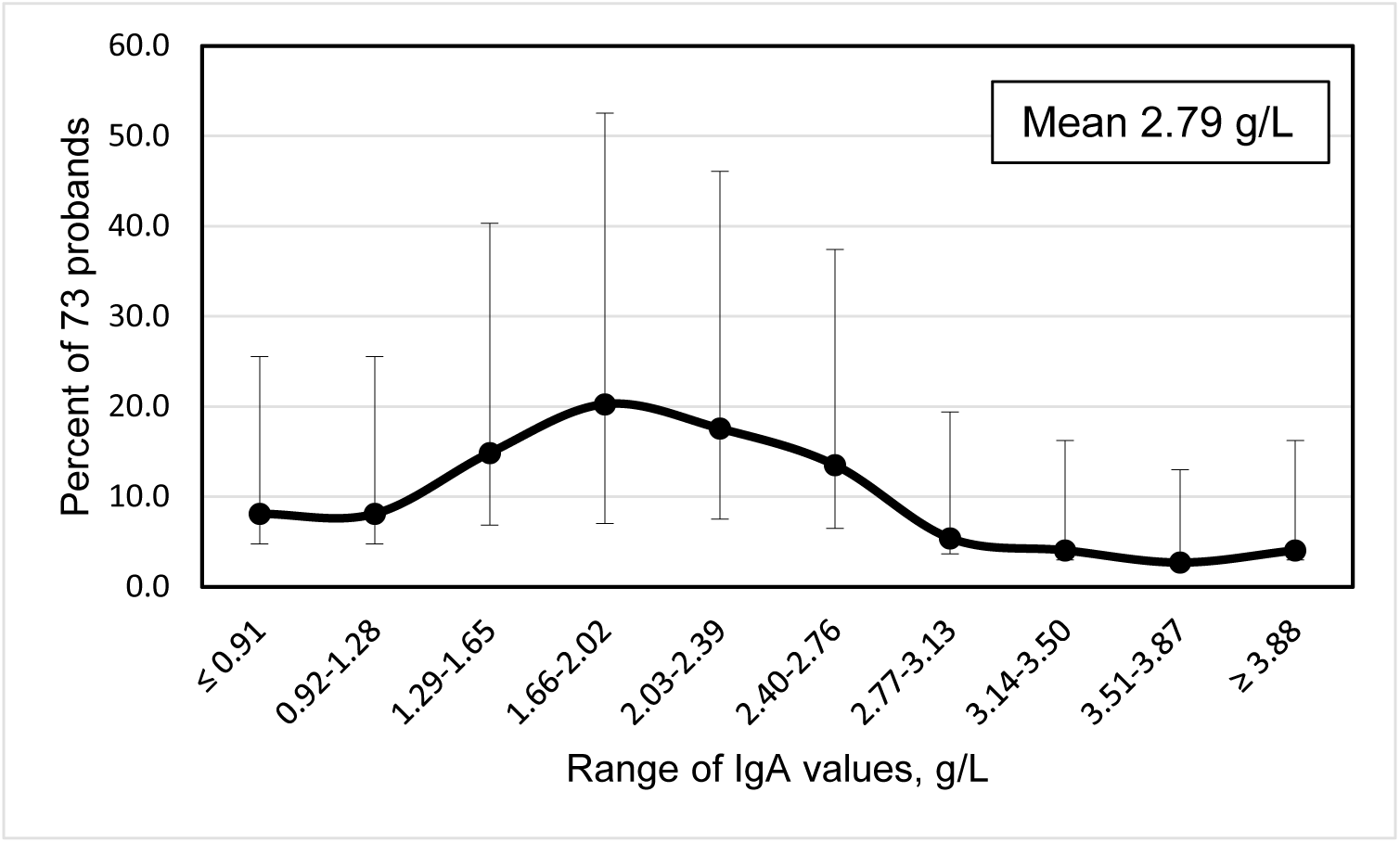
Smoothed frequency distribution of serum IgA levels of 73 hemochromatosis probands with *HFE* p.C282Y homozygosity. Error bars represent 95% confidence intervals of proband percentages.

### Frequency distribution of groups of serum IgM levels

The frequency distribution of groups of IgM levels was skewed to the right (Fig. 2). The range 0.40-1.46 g/L included 71.2% of levels. The range of IgM levels was 0.11-5.13 g/L.

**Figure 2.**
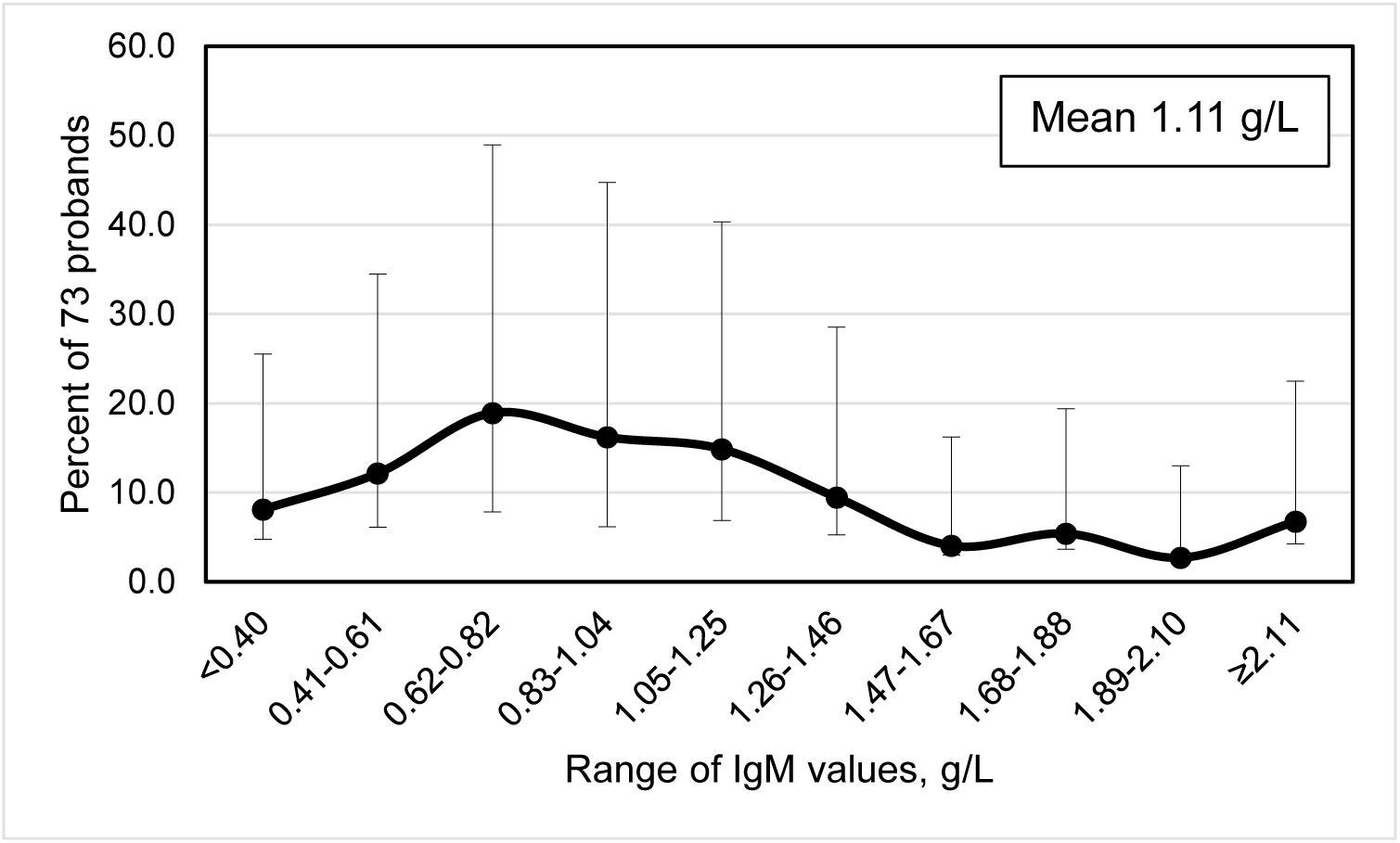
Smoothed frequency distribution of serum IgM levels of 73 hemochromatosis probands with *HFE* p.C282Y homozygosity. Error bars represent 95% confidence intervals of proband percentages.

### Serum IgA and IgM percentiles

The IgA 50.0 percentile value was higher in men than women, although the range of IgA levels in 2.5-97.5 percentiles in men was narrower than that of women (Table 2). All IgM percentile values in women were higher than the corresponding IgM percentile values in men (Table 2).

**Table 2.** Serum IgA and IgM percentiles in hemochromatosis probands.

|  | Percentile | 2.5 | 5.0 | 10.0 | 25.0 | 50.0 | 75.0 | 90.0 | 95.0 | 97.5 |
| --- | --- | --- | --- | --- | --- | --- | --- | --- | --- | --- |
| IgA, g/L | Men (n = 36) | 0.95 | 1.14 | 1.37 | 1.65 | 2.06 | 2.57 | 3.22 | 3.42 | 3.82 |
|  | Women (n = 37) | 0.57 | 0.73 | 0.83 | 1.31 | 1.81 | 2.37 | 3.00 | 4.11 | 5.83 |
|  | All probands (n = 73) | 0.54 | 0.79 | 1.10 | 1.54 | 1.98 | 2.53 | 3.28 | 3.62 | 5.88 |
| IgM, g/L | Men (n = 36) | 0.14 | 0.17 | 0.41 | 0.64 | 0.90 | 1.22 | 1.63 | 1.89 | 2.37 |
|  | Women (n = 37) | 0.31 | 0.43 | 0.52 | 0.79 | 1.11 | 1.40 | 2.20 | 2.59 | 2.71 |
|  | All probands (n = 73) | 0.16 | 0.27 | 0.47 | 0.67 | 0.95 | 1.30 | 1.84 | 2.52 | 2.73 |

IgA, immunoglobulin A; IgM, immunoglobulin M.

### Correlation of serum IgA with age

Pearson’s correlation of IgA with age in all probands was not significant (r_73_ = 0.0030; p = 0.643). Likewise, correlations of IgA with age in 36 male probands (r_36_ = 0.1116; p = 0.517) and 37 female probands (r_37_ = 0.0214; p = 0.900) were not significant.

### Correlation of serum IgM with age

There was a negative Pearson’s correlation of IgM with age in all probands (r_73_ = –0.2733; p = 0.019). In men, there was also a negative correlation of IgM with age (r_36_ = –0.3656; p = 0.028). In women, the correlation of IgM with age was not significant (r_37_ = –0.1766; p = 0.296).

### Associations of serum IgA and IgM with alcohol intake

Corresponding mean serum IgA and serum IgM did not differ significantly between probands who did and those who did not report daily alcohol intake (Table 3).

**Table 3.**
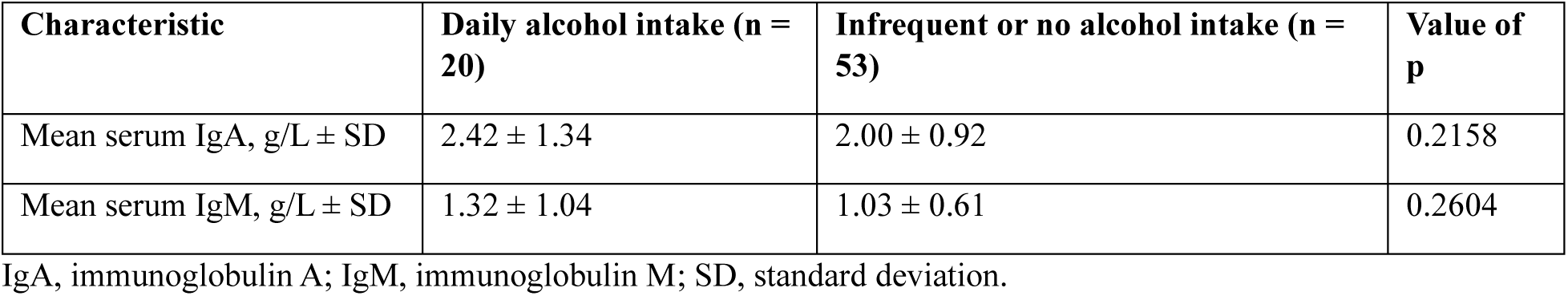
Alcohol intake, IgA, and IgM levels in referred hemochromatosis probands with *HFE* p.C282Y homozygosity.

| Characteristic | Daily alcohol intake (n = 20) | Infrequent or no alcohol intake (n = 53) | Value of p |
| --- | --- | --- | --- |
| Mean serum IgA, g/L ± SD | 2.42 ± 1.34 | 2.00 ± 0.92 | 0.2158 |
| Mean serum IgM, g/L ± SD | 1.32 ± 1.04 | 1.03 ± 0.61 | 0.2604 |

IgA, immunoglobulin A; IgM, immunoglobulin M; SD, standard deviation.

### Associations of serum IgA and IgM with obesity and body mass index

Corresponding mean values of IgA and IgM did not differ significantly between probands with and without obesity (Table 4). Pearson’s correlation of IgA and BMI revealed r_73_ = 0.0548 (p = 0.3428). Correlation of IgM and BMI revealed r_73_ = –0.0984 (p = 0.4042).

**Table 4.**
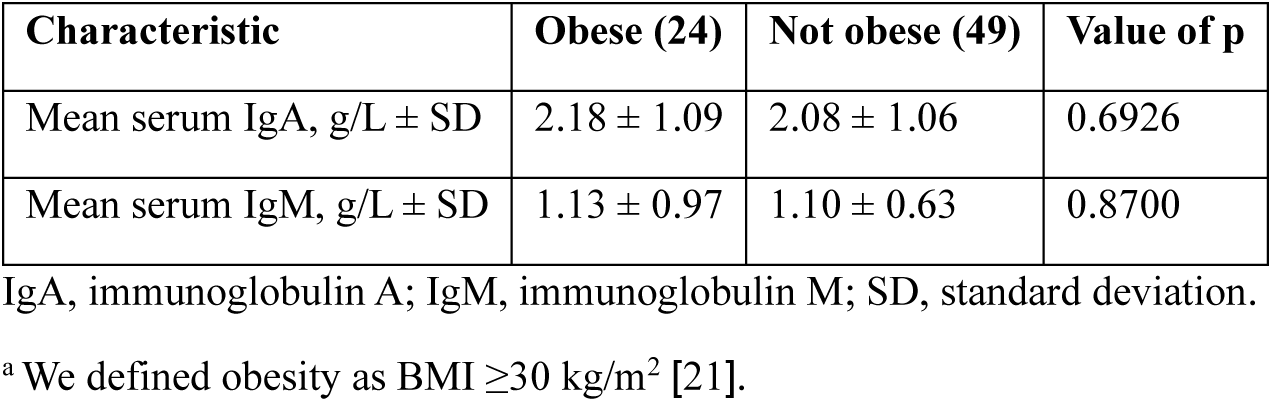
Obesity, IgA, and IgM levels hemochromatosis probands^a^.

| Characteristic | Obese (24) | Not obese (49) | Value of p |
| --- | --- | --- | --- |
| Mean serum IgA, g/L ± SD | 2.18 ± 1.09 | 2.08 ± 1.06 | 0.6926 |
| Mean serum IgM, g/L ± SD | 1.13 ± 0.97 | 1.10 ± 0.63 | 0.8700 |

IgA, immunoglobulin A; IgM, immunoglobulin M; SD, standard deviation.
<sup>a</sup> We defined obesity as BMI $\geq 30$ kg/m<sup>2</sup> [21].

### Regressions on serum IgA and IgM

A final regression on IgA using daily alcohol intake, hand arthropathy, and TS as the independent variables revealed no significant association. A final regression on IgM using male sex, age, SF, and frequency of alcohol intake as independent variables revealed one positive association (daily alcohol intake; p = 0.036) and one negative association (age; p = 0.016). This regression explained 16.1% of the variance of IgM (ANOVA p of regression = 0.017).

### Serum IgA of adults with and without hemochromatosis diagnoses

The mean IgA of 73 hemochromatosis probands and that of 2462 European adults not selected for hemochromatosis diagnoses did not differ significantly (2.11 ± 1.06 g/L vs. 2.16 ± 1.04 g/L, respectively; p = 0.686) (Supplemental Table 1).

The mean IgA of 36 male hemochromatosis probands and that of 918 European men not selected for hemochromatosis diagnoses did not differ significantly (2.19 ± 0.91 g/L vs. 2.15 ± 0.76 g/L, respectively; p = 0.759) (Supplemental Table 2). The mean IgA of 37 female hemochromatosis probands and that of 458 European women not selected for hemochromatosis diagnoses did not differ significantly (2.04 ± 1.20 g/L vs. 1.89 ± 0.66 g/L, respectively; p = 0.219) (Supplemental Table 3).

### Serum IgM of adults with and without hemochromatosis diagnoses

The mean IgM of 73 hemochromatosis probands was lower than that of 1589 European adults not selected for hemochromatosis diagnoses (1.11 ± 0.75 g/L vs. 1.34 ± 0.80 g/L, respectively; p = 0.015) (Supplemental Table 1).

The mean IgM of 36 male hemochromatosis probands was lower than that of 1084 European men not selected for hemochromatosis diagnoses (1.03 ± 0.84 g/L vs. 1.35 ± 0.55 g/L, respectively; p <0.001) (Supplemental Table 4). The mean IgM of 37 female hemochromatosis probands was lower than that of 622 European women not selected for hemochromatosis diagnoses (1.18 g/L ± 0.67 vs. 1.57 ± 0.68, respectively; p <0.001) (Supplemental Table 5).

## Discussion

Novel observations reported in this study include the following: 1) compilation of serum IgA and IgM levels of 73 referred adult hemochromatosis probands with *HFE* p.C282Y homozygosity at diagnosis; 2) determinations of the associations between serum IgA and IgM levels and clinical and other laboratory characteristics of the 73 probands at diagnosis and phlebotomy units removed to achieve iron depletion; and 3) comparisons of mean serum IgA and mean IgM levels of this referral cohort with combined/weighted mean serum IgA and mean serum IgM levels from published cohorts of European adults not selected for hemochromatosis diagnoses or *HFE* genotypes [51–57].

Serum IgA levels of the present probands were not significantly associated with age. In healthy adults affiliated with a North Carolina university, changes in serum IgA after maturity were not significant [58]. In Belgian employees aged 20-65 y, serum IgA levels were not significantly associated with age [52]. In contrast, serum IgA levels increased significantly with age in cohorts of Polish, Italian, Hungarian, and Dutch adults [53,57,59,60]. In a systematic review and meta-analysis, IgA was significantly higher in older than in younger adults [57].

The mean serum IgA levels of the present men and women did not differ significantly, nor did their mean IgA levels differ significantly from those of men and women of European origin whose data we reviewed. The mean IgA of Italian men and women did not differ significantly in two studies [53,54], although the mean IgA of Spanish men was significantly higher than that of Spanish women [55]. In a systematic review and meta-analysis, IgA was significantly higher in men than women [57]. In a study of 93 pairs of monozygotic twins and their spouses and offspring, non-genetic factors accounted for much of the observed variation in IgA levels [61].

Decreasing serum IgM of the present probands was significantly associated with increasing age after adjustment for other variables. In healthy adults affiliated with a North Carolina university, IgM decreased significantly by the sixth decade [58]. Serum IgM was similar or slightly lower in older than younger Hungarian subjects [60]. In Dutch participants, the correlation of IgM and age was not significant [57]. In a systematic review and meta-analysis, IgM was significantly lower in older than younger adults [57].

Serum IgM levels in the present men and women did not differ significantly. In a systematic review and meta-analysis [57], IgM levels were significantly higher in ostensibly healthy women than in men. Higher IgM levels in women are predominantly due to X-linked gene effects [61,62], although IgM levels in women decrease in the fifth and sixth decades [63]. The mean age of the present women was 52 ± 13 y (range 22-80 y), although the correlation of IgM with age in the present women was not significant.

In this study, serum IgA was not associated with reports of daily alcohol intake, although serum IgM was positively associated with daily alcohol intake after adjustment for other factors. Spanish adults whose alcohol intake was >280 g/week had significantly higher mean serum IgA but not higher mean serum IgM than adults who consumed less alcohol [55].

Obesity was not significantly associated with serum IgA and IgM in univariable comparisons in this study. BMI was not significantly associated with serum IgA and IgM after adjustment for other factors. In Spanish adults, obesity was associated with a mild although significant increase in mean serum IgA in a univariable comparison [55]. Mean serum IgM in subjects with and without obesity in the same study did not differ significantly [55].

The serum IgA and IgM levels in the present probands were not associated with positivity for HLA-A*03, a marker for the hemochromatosis ancestral haplotype [19,45]. These results agree with those of a genome-wide association study of Icelanders and Swedes in which neither IgA nor IgM levels were significantly associated with HLA-A [64].

Iron overload occurred in 83.6% of the present probands, although their serum IgA and IgM levels were not significantly associated with TS, SF, or phlebotomy units of blood removed to achieve iron depletion. Mean serum IgA and IgM levels in Indian children with and without iron-deficiency anemia did not differ significantly [65].

It is widely recognized that the penetrance of high-iron phenotypes in *HFE* p.C282Y homozygotes is low [6,17]. Accordingly, seven of the present patients did not have elevated TS and 12 patients did not have elevated SF. Three of these patients had elevated serum levels of hepatic transaminases, a common abnormality in hemochromatosis [66]. Three other patients had macrocytosis, a laboratory phenotype that has been used to identify p.C282Y homozygotes [67,68]. Two women presented with menorrhagia for which iron phenotype testing was indicated [69]. Two other patients had colonic polyps or carcinoma, risks for which were increased in p.C282Y homozygotes in two studies [70,71] although not in a third study [72]. Autoimmune disorders [73] and lethargy [7] are common in p.C282Y homozygotes. Including p.C282Y homozygotes without elevated TS or SF in the present study is supported further by the present observations that neither IgA and IgM levels were significantly associated with either TS or SF after adjustment for other variables. Likewise, IgG subclass levels were not significantly associated with the independent variables TS or phlebotomy units of blood removed to achieve iron depletion in two cohorts of hemochromatosis probands with p.C282Y homozygosity [74,75].

The mean serum IgM levels of the present men and women were significantly lower than the mean IgM of men and women of European origin whose published data we reviewed. Blood mononuclear cells from treated patients with hemochromatosis secreted significantly less IgM after exposure to pokeweed mitogen in vitro than those from normal control subjects [76]. Together, these observations suggest that undefined factors decrease IgM synthesis, secretion, or half-life in plasma in *HFE* p.C282Y homozygotes.

Strengths of this study include the compilation and analyses of serum IgA and IgM levels of a large cohort of referred adult hemochromatosis probands with *HFE* p.C282Y homozygosity and determinations of the associations between IgA and IgM levels and clinical and other laboratory characteristics of the probands at diagnosis of hemochromatosis and phlebotomy units removed thereafter to achieve iron depletion.

Limitations of this study include a lack of measurements of serum IgA and IgM after iron depletion was achieved with therapeutic phlebotomy. This study does not include IgA and IgM levels of age- and sex-matched adults with *HFE* p.C282Y homozygosity identified in population screening or of adults with *HFE* wt/wt (lack of p.C282Y and p.H63D (rs1799945)). The measurement of serum IgA subclasses, the measurement of IgA or IgM in saliva or other secretions, and the assessment of proband IgA or IgM reactivity to specific antigens were beyond the scope of this study. Our review of previous reports of IgA and IgM levels in white adults of European descent not selected for hemochromatosis was not exhaustive.

Uncertainties of this study are related to previous observations that the mean serum IgA and IgM of healthy young men from different European countries measured in the same laboratories using the same techniques differ significantly [77,78]. Thus, mean serum IgA and IgM of the present probands and other cohorts of Europeans whose data we tabulated may differ also in part due to subject selection, age, genetic or non-genetic factors associated with nationality or ethnicity [77,78], or laboratory methods used to measure serum IgA and IgM [79,80]. The present multiple linear regression analyses also indicate that genetic [61,64] or non-genetic [61,81] factors other than those we studied probably influence the IgA or IgM levels of referred adults with hemochromatosis and *HFE* p.C282Y homozygosity.

## Conclusions

We conclude that serum IgM levels of hemochromatosis probands with *HFE* p.C282Y homozygosity are positively associated with daily alcohol intake and inversely associated with age. Mean IgM levels of male and female probands are lower than those of European men and women not selected for hemochromatosis.

## Data Availability

The dataset for analysis is displayed in a Figshare file [16].
Southern Iron Disorders Center. Dataset: Serum IgA and IgM levels in referred hemochromatosis probands with HFE p.C282Y/p.C282Y. Last update: 2024 August. https://doi.org/10.6084/m9.figshare.26764333.v1.

