## Supplemental Tables 1-5 for "Serum IgA and IgM levels in hemochromatosis probands with *HFE* p.C282Y homozygosity"

**Supporting information**

**Supplemental Tables 1-5: Serum IgA and IgM levels of European adults**

**Serum IgA and IgM levels in hemochromatosis probands with *HFE* p.C282Y homozygosity**

Abbreviated title: Serum IgA and IgM in hemochromatosis

James C. Barton^1,2,3*^, J. Clayborn Barton^2^, Luigi F. Bertoli^2,3^, and Ronald T. Acton^2,4^

^1^ Department of Medicine, University of Alabama at Birmingham, Birmingham, Alabama, USA

^2^ Southern Iron Disorders Center, Birmingham, Alabama, USA

^3^ Department of Medicine, Brookwood Baptist Medical Center, Birmingham, Alabama, USA

^4^ Department of Microbiology, University of Alabama at Birmingham, Birmingham, Alabama, USA

*Corresponding author

**Introduction**

We sought to compare the mean serum immunoglobulin A (IgA) and immunoglobulin M (IgM) levels of a cohort of 73 referred Alabama adults with hemochromatosis and *HFE* p.C282Y homozygosity with those of published cohorts of European adults not selected for hemochromatosis diagnoses or *HFE* genotypes.

**Methods**

We performed computerized and manual searches to identify a convenience sample of published reports of serum IgA and IgM measured in cohorts of more than 50 healthy, "control," or general population European adults aged ≥18 y not selected for hemochromatosis diagnoses or *HFE* genotypes. We selected reports that described laboratory methodology for measurement of serum IgA and IgM and expressed cohort results as mean, standard deviation (SD), and 95% confidence interval (CI). We converted IgA and IgM measures published as IU/L to g/L according to the method of Rowe et al. [1]. We excluded reports in which serum IgA and IgM measurements were expressed as medians.

We used a Cochrane’s formula [2] to combine the Ig-specific n, mean, and SD from multiple published cohorts into descriptors of a single group. We used a two-sample t test and n, mean, and SD from each group to compare Alabama proband data with those of Europeans not selected for hemochromatosis diagnoses or *HFE* genotype. We defined values of p <0.05 as significant.

**Results**

**Supplemental Table 1.** Serum IgA and IgM levels of adults

| **Author (year)**^a^ | **Reference** | **Adults, n** | **Method** | **Mean IgA ± SD, g/L [95% CI]** | **Mean IgM ± SD, g/L [95% CI]** |
| --- | --- | --- | --- | --- | --- |
| Šinkov (1973) | [3] | 60 | radial immunodiffusion | 1.62 ± 0.58 [0.93, 2.75] | 1.11 ± 0.39 [0.35, 1.87] |
| Veys (1973) | [4] | 296 | “linear plate” immunodiffusion | 2.08 ± 1.81 [0.79, 6.31] | 0.74 ± 0.87 [0.30, 2.00] |
| Ghessi (1976) | [5] | 603 | single radial immunodiffusion | 2.36 ± 0.74 [0.91, 3.81] | n.a. |
| Quintiliani (1976) | [6] | 773 | immunodiffusion | 1.83 ± 0.65 [0.55, 3.11] | 1.50 ± 0.65 [0.22, 2.78] |
| Gonzalez-Quintela (2008) | [7] | 460 | chemiluminescent enzyme immunoassay | 2.62 ± 1.19 [0.87, 5.76] | 1.47 ± 0.84 [0.46, 3.86] |
| Puissant-Lubrano (2015) | [8] | 270 | immunoturbidimetry | 2.12 ± 0.77 [0.91, 3.93] | n.a. |
| *Combined European data* | *[3-8]* | *2462 (IgA); 1589 (IgM)* | *-* | *2.16 ± 1.04 [2.12, 2.20]* | *1.34 ± 0.80 [1.30, 1.38]* |
| Present cohort (2025) | - | 73 | nephelometry | 2.11 ± 1.06 [1.87, 2.35] | 1.11 ± 0.75 [0.94, 1.28] |
|  |  |  |  | p = 0.686 | p = 0.015 |

CI, confidence interval; n.a., not available; SD, standard deviation.

^a^ Šinkov: healthy individuals aged 21-50 y from Sofia, Bulgaria (30 men, 30 women); Veys: employees aged 20-65 y of the Post, Telegraph and Telephone Administration, Ghent, Belgium (“P.T.T. new”; 296 employees (numbers of men and women not specified)); Ghessi: 603 healthy blood donors aged 21-65 y from Lombardy, Italy (510 men, 93 women); Quintiliani: healthy blood donors aged 20-59 y in Rome, Italy (408 men, 365 women); Gonzalez-Quintela: Caucasian adults aged 18-92 y (median 54 y) from the general population of A-Estrada, northwestern Spain (203 men, 44.1% men); Puissant-Lubrano: 270 healthy blood donors aged 18-68 y from Toulouse, France (136 men, 134 women); Present cohort: referred hemochromatosis probands with *HFE* p.C282Y homozygosity aged 22-80 y (mean 51 ± 13 y) from Alabama, USA (36 men, 37 women).

**Supplemental Table 2.** Serum IgA levels of men^a^

| **Author (year)**^a^ | **Reference** | **n** | **Method** | **Mean IgA ± SD, g/L [95% CI]** |
| --- | --- | --- | --- | --- |
| Ghessi (1976) | [5] | 510 | single radial immunodiffusion | 2.38 ± 0.73 [0.91, 3.84] |
| Quintiliani (1976) | [6] | 408 | immunodiffusion | 1.86 ± 0.69 [0.51, 3.21] |
| *Combined European data* | *[5,6]* | *918* | *-* | *2.15 ± 0.76 [2.10, 2.20]* |
| Present cohort (2025) | - | 36 | nephelometry | 2.19 ± 0.91 [1.89, 2.49] |
|  |  |  |  | p = 0.759 |

CI, confidence interval; SD, standard deviation.

^a^ Ghessi: healthy blood donors aged 21-65 y from Lombardy, Italy; Quintiliani: healthy blood donors aged 20-59 y in Rome, Italy; Present cohort: referred hemochromatosis probands with *HFE* p.C282Y homozygosity aged 22-80 y (mean 50 ± 13 y) from Alabama, USA. CI, confidence interval.

**Supplemental Table 3.** Serum IgA levels of women^a^

| **Author (year)**^a^ | **Reference** | **n** | **Method** | **Mean IgA ± SD, g/L [95% CI]** |
| --- | --- | --- | --- | --- |
| Ghessi (1976) | [5] | 93 | single radial immunodiffusion | 2.23 ± 0.75 [0.73, 3.74] |
| Quintiliani (1976) | [6] | 365 | immunodiffusion | 1.80 ± 0.60 [0.60, 3.00] |
| *Combined European data* | *[5,6]* | *458* | *-* | *1.89 ± 0.66 [1.83, 1.95]* |
| Present cohort (2025) | - | 37 | nephelometry | 2.04 ± 1.20 [1.65, 2.43] |
|  |  |  |  | p = 0.219 |

CI, confidence interval; SD, standard deviation.

^a^ Ghessi: healthy blood donors aged 21-65 y from Lombardy, Italy; Quintiliani: healthy blood donors aged 20-59 y in Rome, Italy; Present cohort: referred hemochromatosis probands with *HFE* p.C282Y homozygosity aged 22-80 y (mean 50 ± 13 y) from Alabama, USA. CI, confidence interval.

**Supplemental Table 4.** Serum IgM levels of men^a^

| **Author (year)** | **Reference** | **n** | **Method** | **Mean IgM ± SD, g/L [95% CI]** |
| --- | --- | --- | --- | --- |
| Šinkov (1973) | [3] | 30 | radial immunodiffusion | 0.97 ± 0.44 [0.12, 1.83] |
| Ghessi (1976) | [5] | 510 | single radial immunodiffusion | 1.50 ± 0.48 [0.57, 3.94] |
| Quintiliani (1976) | [6] | 408 | immunodiffusion | 1.33 ± 0.57 [0.26, 2.44] |
| Puissant-Lubrano (2015) | [8] | 136 | immunoturbidimetry | 0.90 ± 0.49 [0.32, 2.23] |
| *Combined European data* | *[3,5,6,8]* | *1084* | *-* | *1.35 ± 0.55 [1.32, 1.38]* |
| Present cohort (2025) | - | 36 | nephelometry | 1.03 ± 0.84 [0.76, 1.30] |
|  |  |  |  | p <0.001 |

CI, confidence interval; SD, standard deviation.

^a^ Šinkov: healthy individuals aged 21-50 y from Sofia, Bulgaria; Ghessi: healthy blood donors aged 21-65 y from Lombardy, Italy; Quintiliani: healthy blood donors aged 20-59 y in Rome, Italy; Puissant-Lubrano: healthy blood donors aged 18-68 y from Toulouse, France; Present cohort: referred hemochromatosis probands with *HFE* p.C282Y homozygosity aged 22-80 y (mean 50 ± 13 y) from Alabama, USA.

**Supplemental Table 5.** Serum IgM levels of women^a^

| **Author (year)**^a^ | **Reference** | **n** | **Method** | **Mean IgM ± SD, g/L [95% CI]** |
| --- | --- | --- | --- | --- |
| Šinkov (1973) | [3] | 30 | radial immunodiffusion | 1.25 ± 0.29 [0.9, 1.82] |
| Ghessi (1976) | [5] | 93 | single radial immunodiffusion | 1.89 ± 0.49 [0.71, 5.05] |
| Quintiliani (1976) | [6] | 365 | immunodiffusion | 1.69 ± 0.69 [0.33, 3.04] |
| Puissant-Lubrano (2015) | [8] | 134 | immunoturbidimetry | 1.10 ± 0.54 [0.50, 2.60] |
| *Combined European data* | *[3,5,6,8]* | *622* | *-* | *1.57 ± 0.68 (1.52, 1.62)* |
| Present cohort (2025) | - | 37 | nephelometry | 1.18 ± 0.67 [0.96, 1.40] |
|  |  |  |  | p <0.001 |

CI, confidence interval; SD, standard deviation.

^a^ Šinkov: healthy individuals aged 21-50 y from Sofia, Bulgaria; Ghessi: healthy blood donors aged 21-65 y from Lombardy, Italy; Quintiliani: healthy blood donors aged 20-59 y in Rome, Italy; Puissant-Lubrano: healthy blood donors aged 18-68 y from Toulouse, France; Present cohort: referred hemochromatosis probands with *HFE* p.C282Y homozygosity aged 22-80 y (mean 50 ± 13 y) from Alabama, USA.
**References for Supporting Tables**
